# Classifying autism in a clinical population based on motion synchrony: a proof-of-concept study using real-life diagnostic interviews

**DOI:** 10.1101/2023.07.27.23293186

**Authors:** Jana Christina Koehler, Mark Sen Dong, Da-Yea Song, Guiyoung Bong, Nikolaos Koutsouleris, Heejeong Yoo, Christine M. Falter-Wagner

**Author notes:** Corresponding author: Jana C. Koehler. equally contributing first authors (Jana Christina Koehler, Mark Sen Dong). equally contributing senior authors (Heejeong Yoo, Christine M. Falter-Wagner).

## Abstract

Predictive modeling strategies are increasingly studied as a means to overcome clinical bottlenecks in the diagnostic classification of autism spectrum disorder. However, while some findings are promising in the light of diagnostic marker research, many of these approaches lack the scalability for adequate and effective translation to everyday clinical practice. In this study, our aim was to explore the use of objective computer vision video analysis of real-world autism diagnostic interviews in a clinical sample of children and adolescents to predict diagnosis. Specifically, we trained a support vector machine learning model on interpersonal synchrony data recorded in Autism Diagnostic Observation Schedule (ADOS) interviews of patient-clinician dyads. Our model was able to classify dyads involving an autistic patient (n=56) with a balanced accuracy of 63.4% against dyads including a patient with other psychiatric diagnoses (n=38). Further analyses revealed no significant associations between our classification metrics with clinical ratings. We argue that, given the above-chance performance of our classifier in a highly heterogeneous sample both in age and diagnosis, with few adjustments this highly scalable approach presents a viable route for future diagnostic marker research in autism.

## 1 Background

Autism spectrum disorder is characterized by symptoms in social interaction and communication as well as repetitive behaviors. Typically diagnosed during childhood [1], autism is increasingly diagnosed in adulthood over the past years [2], with prevalence estimates around 1% [3]. Due to the lack of clear diagnostic markers, the current gold-standard diagnostic process requires multiple assessments with a trained interdisciplinary clinical team [4], including a diagnostic observation (e.g., Autism Diagnostic Observation Schedule, ADOS-2 [5]), neuropsychological tests, and an interview with a caregiver about the early developmental history (e.g., Autism Diagnostic Interview, ADI-R [6]). While thorough assessments are vital for correct diagnosis, the process itself is lengthy and resource-heavy, causing long waiting times which, thus, comes at a great cost for all involved. Due to the rising demand for diagnostics in recent years, attempts are increasingly made to advance the diagnostic process through personalized prediction approaches based on computational methods such as machine learning. One approach that naturally lends itself to further investigation is the data-driven investigation of existing diagnostic tools such as ADOS. Several studies have been conducted to improve the existing diagnostic algorithm by filtering out a subset of key items predictive for diagnosis. For example, using feature selection-based machine learning on a large data set of children’s ADOS results, Kosmicki et al. [7] significantly reduced the number of relevant items for accurate diagnostic prediction by more than 55%. Küpper and colleagues [8] found that diagnostic prediction performance for adolescents and adults with only five ADOS items was comparable to the originally proposed 11-item diagnostic algorithm.

Nevertheless, this approach is prone to a certain circularity, given the outcome criterion, that is the clinical diagnosis of ASD, is heavily influenced by the features used for prediction [8]. Thus, using machine learning on objective and rater-independent datasets for the screening of potential markers is desirable. Hence, several studies investigated structural or functional brain abnormalities as predictive markers in ASD [9], with promising accuracies especially for younger children [10]. However, methods such as magnetic resonance imaging lack scalability and are impractical to implement in standardized clinical practice. Additionally, those approaches pose special challenges for a sensory-sensitive study population such as autistic individuals. Thus, a more translational approach uses machine learning for diagnostic classification in ASD through digitally assisted diagnostics or digital phenotyping [11], which directly taps the symptomatic behavior. This approach combines the advantages of moving away from the human coding of behaviors while using more scalable methods such as tablet-based movement data or video analysis via computer vision techniques. For instance, Anzulewicz and colleagues [12] reported that a machine learning model trained to identify children with ASD based on their tablet-recorded motion trajectories performed with an accuracy of 93%. In a recent study, Jin et al. [13] developed a pipeline to objectively extract movement features correlated with clinicians’ ratings from children during ADOS interviews.

Although autism is commonly referred to as a disorder of social interaction, thus, implying a certain degree of reciprocity, this aspect is challenging to assess objectively. The increasingly studied phenomenon of reduced interpersonal synchrony in ASD [14] provides such an opportunity. In a previous study [15], we found reduced interpersonal synchrony as derived from motion energy analysis (MEA [16]) in diagnostic interviews with autistic adults as compared to those who did not subsequently receive an autism diagnosis. Furthermore, we explored the predictiveness of interpersonal synchrony between autistic and non-autistic interactants on multiple modalities, finding high accuracy for the synchrony of facial and head movements [17]. However, these studies were conducted with adults, and while motor difficulties in autism tend to persist throughout adulthood [18], little is known about the predictive power of synchrony alterations in children.

In a study on video-based pose estimation, Kojovic et al. [19] investigated videos of ADOS interviews with small children. Their deep neural network analysis of multiple aspects of non-verbal interaction differentiated between autistic children and typically-developing (TD) children with an accuracy of 80.9% and additionally revealed associations between their model and the overall level of symptomatology. Thus, modeling based on direct extraction of predictive features from diagnostic videos opens a promising avenue for the clinical setting.

Our aim in this proof-of-concept study was to investigate automatic video analysis as a scalable approach to screen for synchrony alterations as an objective marker to classify autism in children and adolescents. To this end, we trained several support vector machine (SVM) classification models using synchrony features extracted from videos of real-life ADOS interviews and investigated the associations of our classifiers’ outputs with professional clinical ratings.

Importantly, to explore model specificity in a realistic clinical scenario, we used a representative clinical sample that included participants who were subsequently diagnosed with ASD as well as patients with other psychiatric diagnoses.

## 2 Methods

In the following, we report the details of our prediction model following the Transparent Reporting of a Multivariable Prediction Model for Individual Prognosis or Diagnosis (TRIPOD) guidelines [20].

### 2.1 Sample

The ADOS videos and their related datasets were compiled from two different sources at Seoul National University Bundang Hospital: the patient pool of the psychiatric outpatient clinic for children and adolescents as well as from a study population of an unrelated study that included the ADOS. Therefore, the inclusion criteria and available data slightly differed. Patients referred to the outpatient clinic underwent extensive clinical examination to evaluate the presence of an ASD or differential diagnosis. Additional information on comorbidities and medication for this subsample is available in the supplementary material (see Supplementary Table S4.1) and was not included in the final analysis. For the patients from the unrelated study, ADOS was performed as part of the study protocol, though the diagnosis had either already been suspected or given elsewhere. In contrast to the outpatient pool, exclusion criteria were applied in the unrelated study which comprised severe motor impairments restricting patients from engaging in the required ADOS activities, as well as sensory-related issues or selective mutism. No age limit applied.

For all cases from both sources, the autism diagnosis was confirmed as a best clinical estimate consensus diagnosis by two psychiatrists, taking into account ADOS and ADI-R results, as well as other neuropsychological assessments.

An overview of the current sample compilation procedure can be found in Figure 1. All available ADOS video materials were initially screened for the first occurrence of at least five minutes of consecutive and unobstructed footage for every participant based upon the following criteria: (a) steady camera position and constant lighting, (b) camera angle that includes the head and upper body of both participant and ADOS administrator, (c) both participant and administrator being seated throughout all video frames (i.e., no freeplay, no running around), (d) and no use of props. As only ADOS modules three and four include longer instances of free-flowing conversation, the final sample was comprised of these modules. Excerpts were taken from the tasks *Emotion*, *Conversation and Reporting*, *Social Difficulties and Annoyance*, *Job/School Life*, *Friends, Relationships, and Marriage*, and *Loneliness*. Due to the semi-structured nature of ADOS, the final clips differed in length, ranging from 5:15 minutes to 14:37 minutes (mean length = 7:20 minutes). Interviews were conducted by six different administrators. All videos had a frame rate of 29.95 seconds.

**Figure 1.**
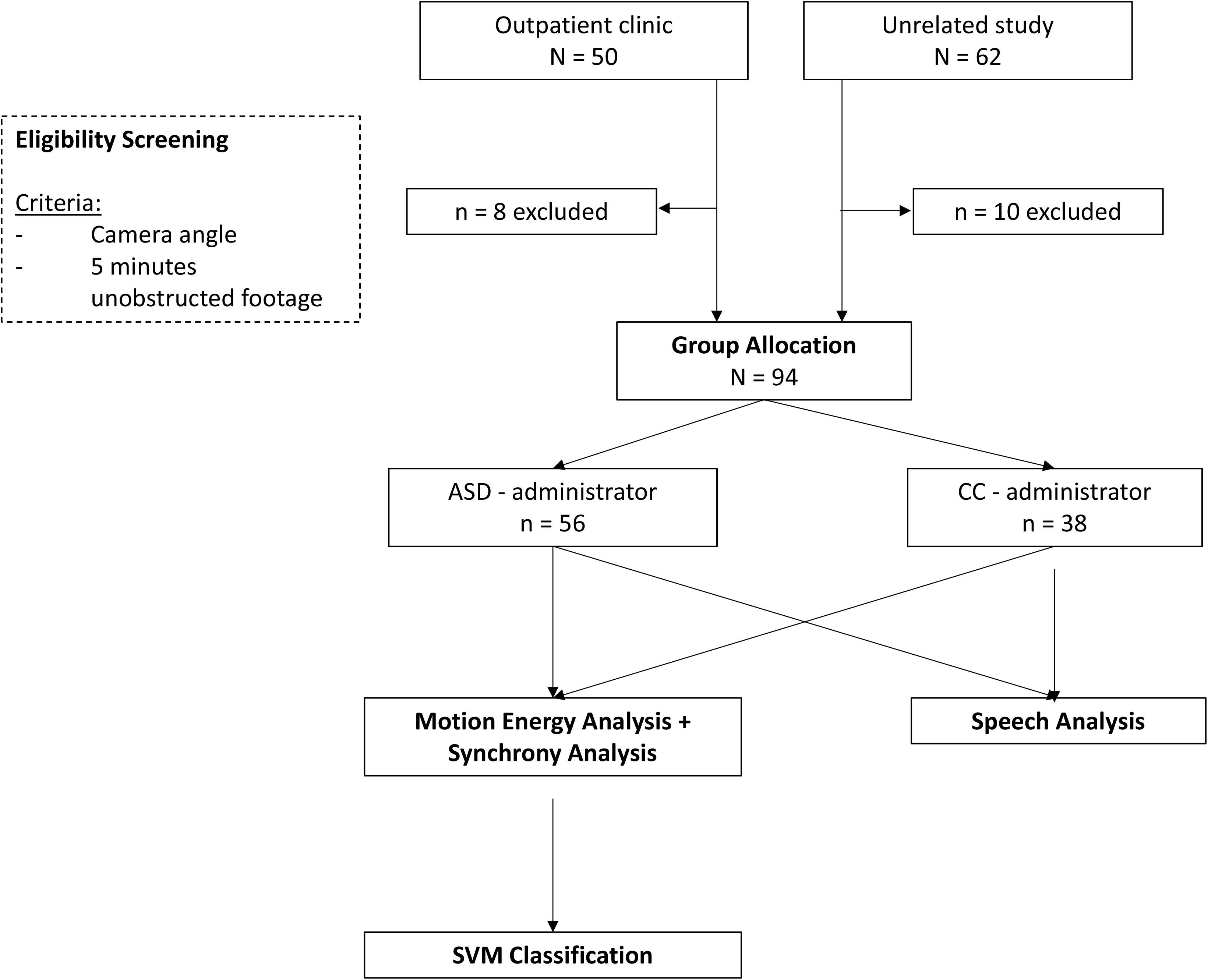
Consort chart of the current sample compilation.

The final dataset consisted of 56 participants with a diagnosis of ASD and 38 participants with other psychiatric conditions (i.e., n = 4 Intellectual Disability, n = 1 Developmental Delay, n = 10 ADHD, n = 1 Tourette Syndrome, n = 4 Depressive Disorder, n = 1 Social Phobia, n = 1 Anxiety Disorder, n = 2 Bipolar Disorder) or within the wider autism phenotype (n = 2), as well as n = 12 typically-developing (TD) children (including 8 unaffected siblings). This resulted in two diagnostic group allocations: ASD-administrator or clinical control (CC)-administrator.

The authors assert that all procedures contributing to this work comply with the ethical standards of the relevant national and institutional committees on human experimentation and with the Helsinki Declaration of 1975, as revised in 2008. The study to use fully anonymized data collected retrospectively and prospectively were approved by the Institutional Review Board at Seoul National University Bundang Hospital (IRB no., B-1812-513-105; B-1912-580-304). Informed consent was obtained for the participant data collected prospectively from both participants and, in case the participant was a minor, their parent or legal guardian. A separate informed consent for the analysis of completely anonymized retrospective data was waived.

### 2.2 Video pre-processing and synchrony computation

Motion Energy Analysis (MEA) [16] was applied to all video clips, defining two regions of interest per participant and administrator (head and upper body). MEA extracts frame-to-frame gray-scale pixel changes. Keeping the camera position, lighting and background constant, all pixel changes above a manually set threshold represent movement within the regions of interest. After careful visual inspection of the resulting data quality, a threshold of eight was chosen for all videos.

Raw motion energy time series were subsequently forwarded to pre-processing using the RStudio package rMEA [21]. Videos were filmed in four different rooms. To account for potential biases of any distortions in the video signals, all MEA time series were scaled by standard deviation and smoothed with a moving average of 0.5 seconds, according to the default setting in rMEA. A comparison analysis of potential feature differences depending on the room can be found in the supplementary material (S2.2).

Interpersonal synchrony between participant and administrator in their head and body motion was computed with windowed cross-lagged correlations. In line with a previous analysis of diagnostic interviews with autistic adults [15], a window size of 60 seconds was chosen. To capture all instances of synchrony, time series were cross correlated with lags of 5 seconds and increments of 30 seconds. All values in the resulting cross-correlation matrices were converted to absolute Fisher Z values. Time series were subsequently shuffled and randomly paired into 500 pseudodyads. Cross correlations were conducted in the same manner, yielding a measure of pseudosynchrony per artificial dyad. They were subsequently compared to the interpersonal synchrony values to assess whether the interpersonal synchrony values were above-chance. Detailed results can be found in the supplementary material (S2.3).

Moreover, following procedures from Georgescu et al. [22], intrapersonal head and body coordination was computed for every patient, using window sizes of 30 seconds, lags of 5 seconds and a step size of 15 seconds.

Lastly, we derived the head and body movement quantity per participant from the respective MEA time series. Following previous procedures [15,23], they were defined as the number of frames with changes in motion energy divided by the total number of frames, resulting in four values per dyad (two for participant and administrator, respectively).

In addition to the processing of motion, we submitted our videos to an exploratory vocal output analysis. For this purpose, the audio tracks of the selected clips were processed with the software Praat [24] to semi-automatically extract annotations of intervals of vocalizations and silences. As there was no speaker distinction within the audio tracks, this analysis was considered exploratory and is not included in the main machine learning analysis. Details can be found in the supplementary material (S1.1).

### 2.3 Feature engineering

Because the videos in our sample varied in both length and conversational content (see Supplementary Material S2.1), as well as to account for the interview context, our aim was to gain the best estimate of the overall synchrony (i.e., instances in and out of synchrony). For this reason, summary statistics of each cross-correlation matrix were computed (i.e., minimum, maximum, mean, median, standard deviation, skew, and kurtosis), resulting in seven features per participant-administrator dyad and region of interest (ROI). We additionally computed the same summary statistics for the intrapersonal head-body coordination of each participant. This approach slightly differed from a previous study [17], where we were interested in the trajectory of maximum synchrony instances during naturalistic and free-flowing conversations. To comply with previous procedures, we additionally computed a feature set using a peak-picking algorithm to obtain a measure of the trajectory of the highest synchrony instances during each interview. Details and results can be found in the supplementary material (S2.4).

The final feature set for each dyad consisted of 25 features per participant-administrator dyad (see Supplementary Table S4.2): seven interpersonal synchrony features per dyad and ROI (head and body), seven features for the intrapersonal head-body coordination of every participant, as well as four features for the individual amount of head and body movement of both interactants. IQ and sex of the participant were additionally included as features in a second model, as both are frequently associated with autism symptomatology and the likelihood of receiving a diagnosis [25,26].

### 2.4 Support Vector Machine (SVM) Learning Analyses

We trained two separate binary machine learning models to classify between dyad type: (1) a “behavioral” model containing only synchrony data objectively extracted from the videos (MEA), and (2) a model additionally containing sex and IQ as demographic features (MEA + DEMO). Age was regressed out in both models. By constructing two separate models, we could explore whether demographic features frequently associated with ASD might improve the purely behavioral predictive performance. A L1-loss LIBSVM algorithm was chosen for both models, as it is frequently used in psychiatric research [27], known to perform robustly with reduced sample sizes [28]. In each model, the SVM algorithm independently modeled a linear relationship between features and classification labels by optimizing a linear hyperplane in a high-dimensional feature space to maximize separability between the dyads. Subsequently, the data was projected into the linear kernel space and their geometric distance to the decision boundary was measured. Thus, every dyad was assigned a predicted classification label and a decision score.

Machine learning analyses were conducted in NeuroMiner (Version 1.1; https://github.molgen.mpg.de/pages/LMU-Neurodiagnostic-Applications/NeuroMiner.io/) [29], an open-source mixed MATLAB [30]-Python-based machine learning library. To prevent any possibility of information leakage between training and testing data, our diagnostic classifiers were cross-validated in a repeated, nested, stratified cross-validation scheme. We used ten folds and ten permutations in the outer CV loop (CV2) and ten folds and one permutation in the inner loop (CV1). Specifically, at the CV2 level, we iteratively held back 9 or 10 participant-administrator dyads as test samples, while the rest of the data entered the CV1 cycle, where the data were again split into training and validation sets. This way training and testing data were strictly separated, with hyper-parameter tuning happening entirely within the inner loop while the outer loop was exclusively used to measure the classifier’s generalizability to unseen data and generate decision scores for each dyad in this partition. This process was repeated for the remaining folds, after which the participants were reshuffled within their group and the process was repeated nine times, producing 10×10=100 decision scores for each held out participant. The final median decision score of each held out dyad was computed from the scores provided by the ensemble of models in which given dyad had not been used at the CV1 level for training or hyperparameter optimization. Additionally, the stratified design ensured that the proportion of the diagnostic groups in every fold would adequately reflect the proportion of the diagnostic group in the full sample and, thus, guarantee that each class is equally represented in each test fold to avoid bias during model training. The preprocessing settings for the respective models can be found in Table 1.

**Table 1.** SVM Classification Model Descriptions.

| Model | Features | Preprocessing Pipeline |
| --- | --- | --- |
| MEA | Interpersonal head synchrony (7)<br>Interpersonal body synchrony (7)<br>Intrapersonal head-body coordination of patient (7)<br>Total head and body movement (4) | 1. Scaling between 0 and 1<br>2. Pruning of non-informative features (zero variance, infinite values)<br>3. Age as covariate (partial correlation) |
| MEA + DEMO | Interpersonal head synchrony (7)<br>Interpersonal body synchrony (7)<br>Intrapersonal head-body coordination of patient (7)<br>Total head and body movement (4)<br>IQ (1)<br>Sex (1) | 1. Scaling between 0 and 1<br>2. Pruning of non-informative features (zero variance, infinite values)<br>3. k-nearest neighbor imputation of missing values<br>4. Age as covariate (partial correlation) |

*Note. Number of features of respective modality in parentheses. Missing IQ values (16% of cases) were imputed using k-nearest neighbor imputation.*

Class imbalances were corrected for by hyperplane weighting. Balanced Accuracy (BAC = [sensitivity+specificity]/2) was used as the performance criterion for hyperparameter optimization. The C parameter was optimized in the CV1 cycle using 11 parameters within the following range: 0.0156, 0.0312, 0.0625, 0.1250, 0.2500, 0.5000, 1, 2, 4, 8, and 16, which represent the default settings in NeuroMiner [29]. Model significance was assessed through label permutation testing [31], with a significance level α = .05 and 1000 permutations. The permutation testing procedure determines the statistical significance of a model’s performances (i.e., BAC) by using the current data compared to models trained on the dataset but with the labels randomly permuted. The predictive pattern of the models was extracted using cross-validation ratio (CVR) and sign-based consistency. Firstly, CVR was computed as the mean and standard error of all normalized SVM weight vectors concatenated across the entire nested CV structure. CVR measures pattern element stability and was defined as the sum across CV2 folds of the CV1 median weights divided by their respective CV1 standard error, all of which was subsequently divided by the number of CV2 folds [32]. Secondly, we used the sign-based-consistency method [33] to test the stability of the predictive pattern by examining the consistency of positive and negative signs of the feature weight values across all models in the ensemble (see Supplementary Material S1.2 for additional information). Feature stability was assessed for statistical significance at α = .05, using the Benjamini-Hochberg procedure of false discovery rate correction (FDR) [34].

### 2.5 Associations of SVM model and clinical variables

To investigate potential underlying clinical factors associated with our classification models, post-hoc correlation analyses with the SVM decision scores and ADOS, as well as ADI scores were performed in RStudio (version 2022-07.2) [35]. A dyad’s predicted SVM decision score represents their distance from the hyperplane. ADOS scores included domain scores for social affect (SA) and restricted and repetitive behaviors (RRB), as well as the total score (Total). Because our sample included data from both modules three and four, calibrated severity scores [36,37] were used for the correlation analyses for better comparison. For ADI-R, ratings on three subdomains based on caregiver report were used: reciprocal social interaction (A), social communication (B), and restricted and repetitive behaviors (C). Statistical significance was determined at α = .05 and two-sided *p* values were corrected for multiple comparisons using FDR.

### 2.6 Exploratory SVM analysis

To further address the specificity of synchrony, given that phenotypic movement difficulties overlap in neurodevelopmental disorders (e.g., dyspraxia and autism, or hyperkinetic movement in ADHD), the MEA classifier was retrained within the same sample but using different class labels: i) a neurodevelopmental disorders class, which grouped all 74 patients with a diagnosis of a neurodevelopmental disorder as defined by DSM-5 [38] (n = 56 ASD, n = 10 ADHD, n = 1 Developmental Delay, n = 1 Tourette Syndrome, n = 4 Intellectual Disability, n = 2 Broad Spectrum/Pervasive Developmental Disorder – Not Otherwise Specified (PDD-NOS)), and ii) a clinical control group consisting of the 20 patients with other psychiatric diagnoses or typically-developing participants (n = 12 TD including 8 unaffected siblings, n = 1 Anxiety Disorder, n = 2 Bipolar Disorder, n = 4 Depressive Disorder, n = 1 Social Phobia). The stratified CV structure was adapted accordingly.

## 3 Results

### 3.1 Sample Description

A description of the final sample grouped according to the ADOS module can be found in Table 2. A chi-square test of independence revealed no significant association between the diagnostic group and sex (χ^2^(1,94) = .045, *p* = .831). Though naturally participants across both modules differed in age, there was no significant difference in age between diagnostic groups within each module. Because final diagnosis was partly based on ADOS and ADI-R results, autistic patients across both modules had significantly higher ADOS as well as ADI-R scores compared with the clinical control group. Best-estimate IQ values were significantly higher in the CC group for module 3. This effect was reversed in module 4, with autistic patients scoring significantly higher on their respective IQ assessment.

**Table 2.**
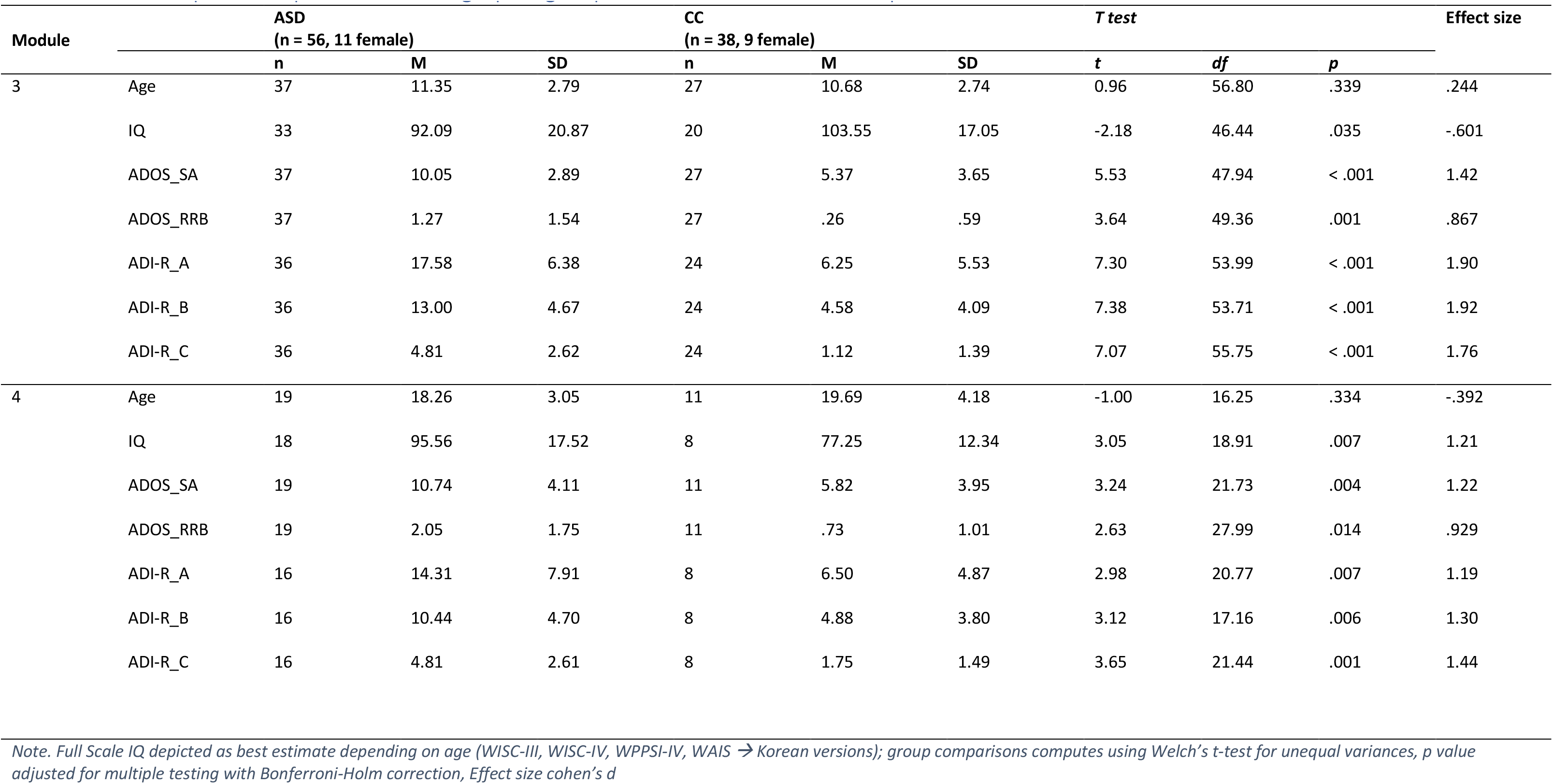
Sample description and demographic group differences across subsamples.

### 3.2 SVM Classification Performance and Feature Importance

Using only motion energy analysis data and regressing out age, our MEA model was able to classify interview dyads with an autistic participant as opposed to those with other psychiatric diagnoses with a BAC of 63.4% (Figure 2). Detailed performance metrics, i.e., sensitivity, specificity, accuracy, positive and negative predictive values, and Area-Under-the-Receiver-Operating-Curve (AUC) can be found in Table 3. There was no significant residual association between age (M = 13.53, SD = 4.70) and the model’s resulting decision scores (M = .19, SD = .89) after regressing out age during pre-processing (*r_Pearson_* = .06, *p* = .558). The model that additionally included sex and IQ as features (MEA + DEMO) had a lower BAC of 59.4% (Sensitivity = 71.4%, Specificity = 47.4%, AUC = .58[CI = .46 - .70], also see S4.3 Supplementary Table).

**Figure 2.**
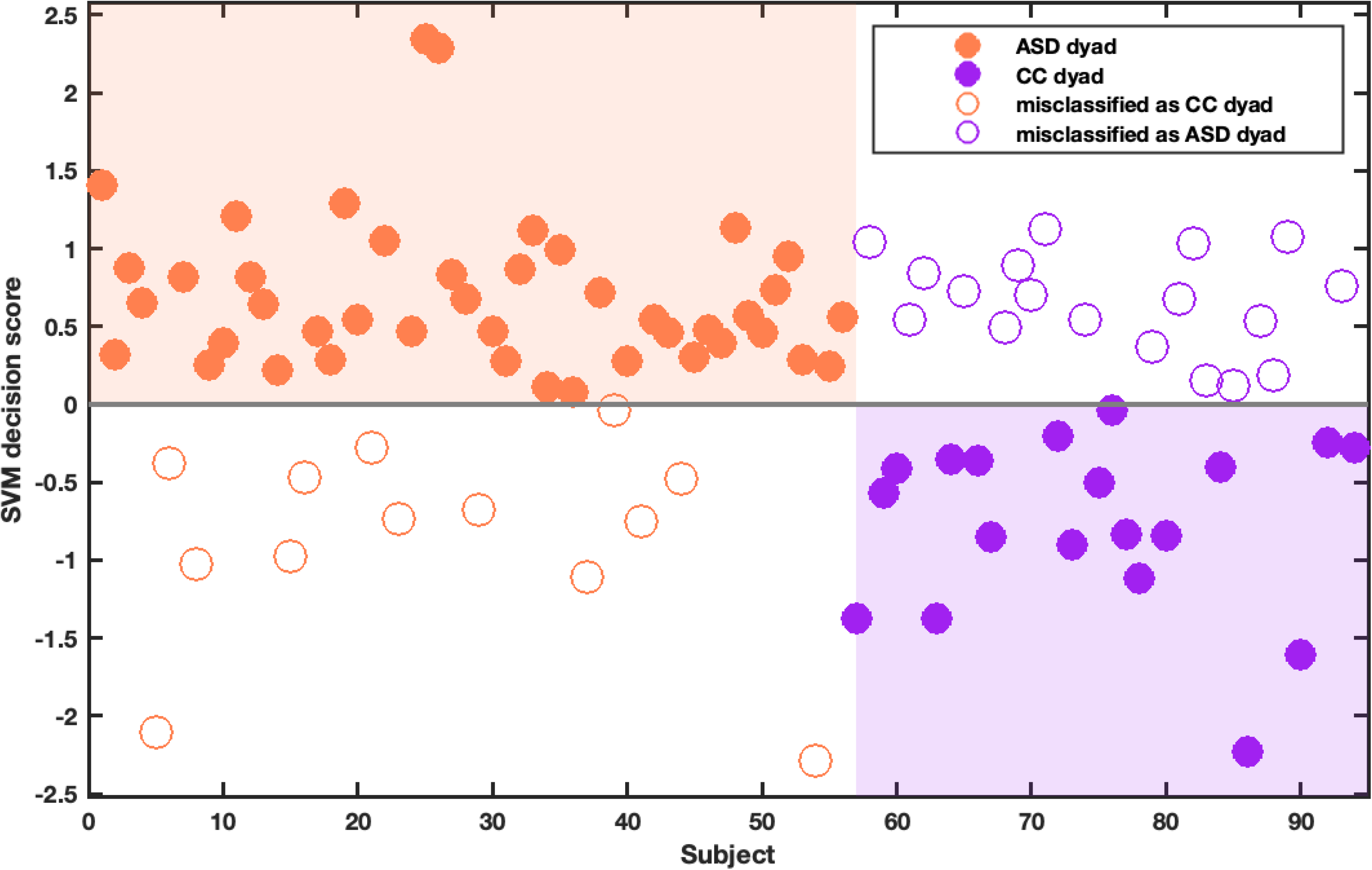
SVM classification results of ASD vs. CC patient-administrator dyads *Note. Figure depicts mean classifier scores of each dyad in the model containing only MEA data, resulting in a balanced classification accuracy of 63.4%. The further the score is from the decision boundary, the more likely this dyad was predicted as belonging to their respective class*.

**Table 3.**
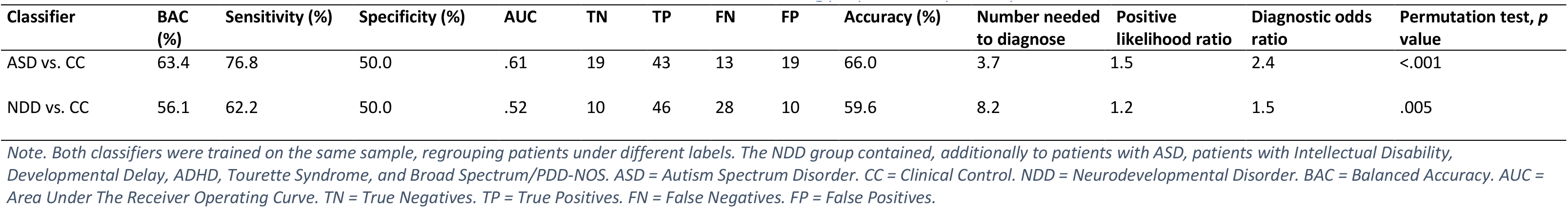
Classification metrics for SVM classifiers based on Motion Energy Synchrony Analyses between Patient and Administrator.

A closer investigation of the cross-validation ratio revealed that classification towards the autism-administrator dyads was driven by higher kurtosis and skewness of their body synchrony values (Figure 3a). This means that a dyad with more pronounced outliers in their body synchrony, especially in the positive direction, was considered more autistic. In contrast, our model considered higher mean body synchrony values as non-autistic. Sign-based consistency revealed that this effect was relatively stable (Figure 3b). Interestingly, the opposite effect was visible for head synchrony: higher kurtosis and skewness of head synchrony values were considered non-autistic, whereas higher mean head synchrony values were considered autistic. However, this was not consistent and of less feature importance than body synchrony.

**Figure 3.**
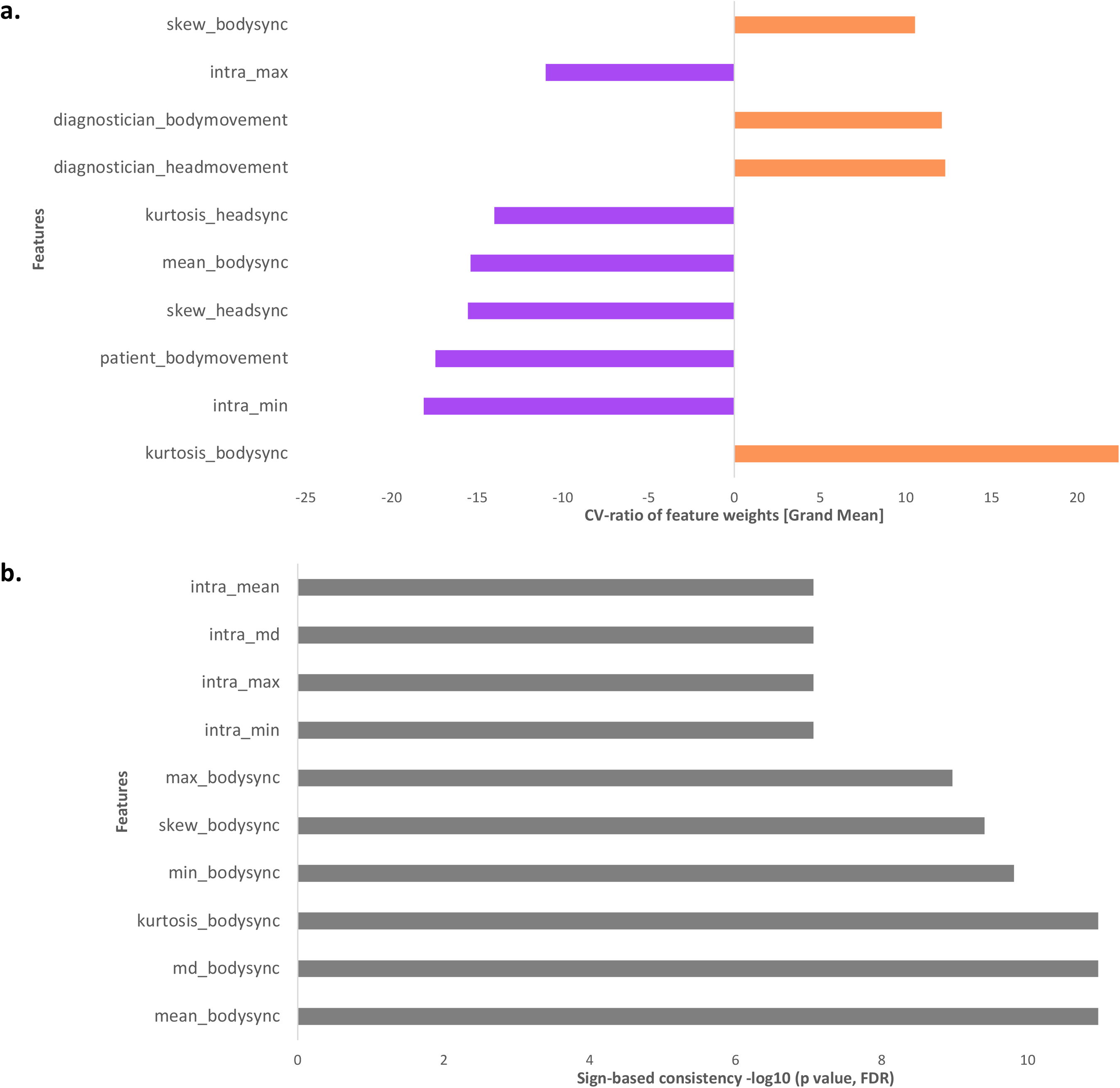
Feature importance of SVM model *Note. Only the ten most important features are depicted. a. Cross-validation ratio. Figure depicts the sum across CV2 folds of the selected CV1 median weights divided by the selected CV1 standard error, which is subsequently divided by the number of CV2 folds. Absolute values >= 2 correspond to p <= .05, absolute values >=3 correspond to p <= .01. b. Sign-based consistency. The importance of each feature was calculated as the number of times that the sign of the feature was consistent. The depicted scores represent the resulting negative logarithm of p values that were corrected using the Bonferroni-Holm false-discovery rate. Sign-based consistency - 10log(p) >= 1.3 is equivalent to p <= .05*.

A closer look at the movement parameters of both participant and administrator revealed that more movement by the administrator was taken into account when classifying an autistic dyad, whereas more movement by the participant was classified as a clinical control dyad.

A comprehensive list of cross-validation ratios and sign-based consistencies for all features of the MEA model can be found in the Supplementary Material (Supplementary Figures 2 and 3).

### 3.3 Associations between SVM model scores and clinical variables

We conducted a range of correlation analyses of the resulting SVM scores of our models with ADOS-2 [5] and ADI-R [6] domain and total scores (Figure 4). ADI-R data was incomplete for ten participants, who were discarded from the respective analysis.

**Figure 4.**
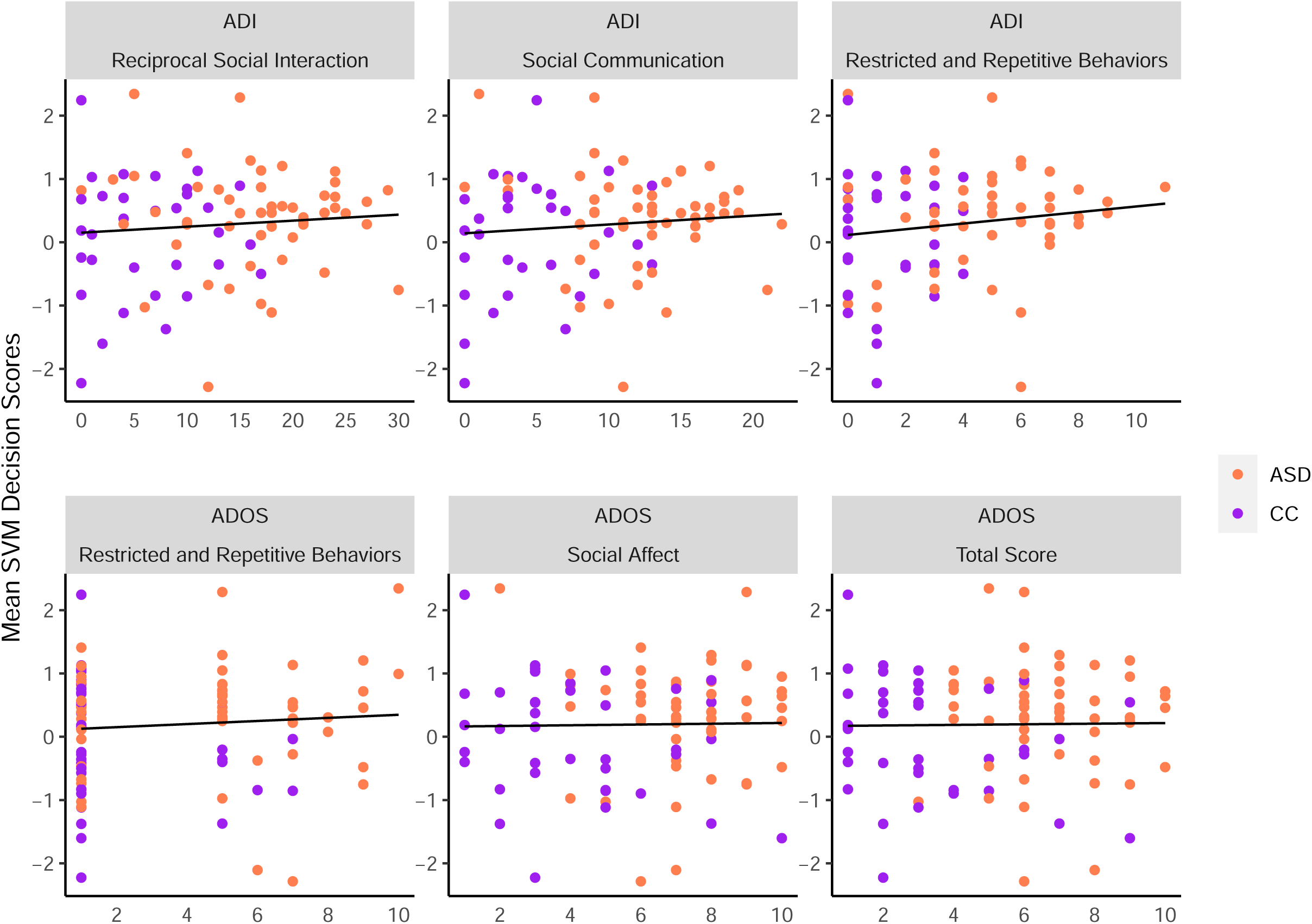
Association between SVM decision scores of MEA classifier and ADI and ADOS domain scores *Note. ADOS scores were transformed to calibrated severity scores following procedures in* [36,37]*. It should be noted that while the initial class labelling was heavily influenced by both ADOS-2 and ADI-R results, nevertheless, they were not sufficient for diagnosis in this sample*.

In general, classification towards the autistic group was loosely associated with higher ADI-R ratings on all three scales, although these findings were not statistically significant. No significant associations were found for the ADOS ratings. Detailed correlation results can be found in S4.4 Supplementary Table.

### 3.4 Exploratory SVM analysis: NDD vs. CC

When regrouping the present sample and classifying participants with neurodevelopmental disorders in general and clinical controls based on motion energy synchrony (analogous to the MEA model), the BAC decreased to 56.1% (Table 3).

## 4 Discussion

This proof-of-concept study aimed to explore the predictability of autism from non-verbal aspects of social interactions between participants and clinicians using videos of real-life diagnostic interviews. Our classification algorithm solely trained on objectively quantified synchrony values was able to predict autism in a representative clinical sample with a BAC of 63.4%. A separate model including demographic features frequently associated with the likelihood of an autism diagnosis (i.e., sex and IQ) yielded a lower balanced accuracy and, thus, did not improve predictive performance. Feature importance analyses revealed the impact of body synchrony and movement quantity for diagnostic classification. Slight but non-significant associations were found with ratings based on parent’s reports (ADI-R), while we did not find any visible associations with ratings by clinicians. When classifying neurodevelopmental disorders in general against other psychiatric diagnoses, accuracy was lower than the base model, possibly suggesting a non-verbal social interaction signature specific to autism.

Compared to Kojovic et al. [19], the accuracy of our classifier based on motion energy synchrony data between participants and administrators was reduced. This might be due to several reasons: First, our sample was heterogeneous in terms of diagnosis and age. Instead of classifying ASD against TD children, our classifier was trained on a real-life clinical sample, including a range of diagnoses often co-occurring in autism. Reduced interpersonal synchrony has been reported for adults with other psychiatric diagnoses such as depression [39] and schizophrenia [40]; the former being a frequent co-occurring condition in ASD [41] and the latter sharing phenomenological overlaps with autism [42]. For the sake of completeness, we included information on comorbidities and medication in the supplementary material. However, due to the limited availability of this information for many participants, we did not run any analyses on these data. Future studies should investigate the influence of co-occurring and differential diagnoses by, e.g., running clustering analyses. We controlled for the large age range (5.5 – 28.7 years) present in our sample by including chronological age as a covariate, leaving no significant residual association of the model’s decision scores with age. However, while reduced interpersonal synchrony has been found across the lifespan of individuals on the autism spectrum [14], they have yet to be investigated in direct comparison and the association to general motor skills remains unclear. In our sample, the continuing development of motor skills with age could have resulted in larger heterogeneity of the ability to synchronize and reduced classification performance. Another approach to increase classification performance could incorporate multi-modal aspects of synchrony. In the present study, we focused on head and body motion synchrony. However, previous research has shown high predictability of, e.g., facial expression synchrony [43]. In fact, we previously found that facial expression synchrony between two adults was superior to body movement synchrony in predicting autism [17]. As our videos were filmed from a side perspective, the automated analysis of facial expression with current algorithms requiring the presence of certain facial key points was not possible. However, slight changes in the setup, i.e., including frontal recording of distinct facial movements, could possibly improve predictive performance in the future. Additionally, the synchronization of speech and vocal output in interactions has been found to be reduced in autism [44,45]; although, the generalizability of vocal markers across studies is rather limited as suggested by a recent investigation [46]. Furthermore, closer investigation of more fine-grained non-verbal aspects of social interaction provides the distinct advantage such that markers across the entire spectrum could be explored, given that an estimated 25% of individuals on the autism spectrum are non-verbal [47]. Thus, the approach presented in this study is straightforward to adapt for this purpose.

When closely assessing the feature weights, we found that the classification was driven by body synchrony and the clinician’s total amount of body movement. More specifically, classification towards the autistic group was driven by greater movement by the administrator, while more participant movement was associated with classification towards the clinical control group. As MEA is a measure of motion energy rather than a measure of movement quality, this might possibly also reflect a unique feature of the diagnostic interview context, i.e., the clinician documenting on a clipboard and tending to document more meticulously if a patient exhibited more conspicuous behaviors. In contrast, our clinical control group included patients with attention deficit hyperactivity disorder (ADHD), a diagnosis commonly associated with elevated movement [38]. While this suggests a tendency of our model to classify movement, rather than synchrony, definite interpretations of the feature weights should be exhibited with caution before being validated on a larger sample.

Contrary to Kojovic and colleagues [19], we could not detect significant associations between our classifier based on synchrony data and clinical variables in our sample. This could be due to the differences of sample characteristics between both studies. Importantly, the former study classified children with autism against TD children. In clinical outpatient units the representative comparison group is heterogeneous concerning differential diagnoses. As such, our comparison group was more heterogenous with regard to diagnosis as it included children with other psychiatric diagnoses or social communication difficulties. Decreased specificity of ADOS in populations more representative of the real-world clinical setting has been reported in previous studies [48,49]. This was also visible in the overlap of ADOS and ADI severity scores between both groups in our sample. In an exploratory analysis to increase accuracy, we employed a SVM classification on a re-labelled sample, grouping ASD with other neurodevelopmental disorders as defined by the DSM-5 [38]. However, this model performed slightly above chance, suggesting a synchrony signature specific to autism. Yet, we recognize that this finding needs external validation in order to be further interpreted.

Our study has several limitations that should be considered: First, the videos analyzed in this study were not initially recorded for the purpose of automated machine learning-based analysis procedures. For this reason, the setup varied regarding background and camera angles depending on the different rooms. This could also have contributed to the lack of significant differences in our comparison to pseudo-synchrony (see Supplementary materials S2.3). However, we consider this a feature, rather than a flaw of our approach. When comparing the synchrony values between the different rooms, we could not detect significant differences, underling the scalability of our setup. This is in line with Kojovic and colleagues [19] who investigated their computer vision algorithm with two validation samples, finding minimal influence of video conditions. However, for future reference, we have compiled recommendations for a more standardized recording protocol of ADOS which can be found in the supplementary material (S3). Additionally, we recommend the use of separate microphones to allow for more elaborate analyses of verbal interaction, as well as the use of cameras for more fine-grained facial expression analyses.

Secondly, because our videos differed in length, the use of summary statistics as best estimate measures of interpersonal synchrony were deemed most suitable. However, this approach cannot capture the temporal dynamics of synchrony throughout a conversation. During free-flowing conversations, interactants tend to move in and out of synchrony over time [50], suggesting a certain flexibility in interpersonal alignment. However, no clear evidence exists regarding interview contexts. Thus, future research should investigate synchrony trajectories in more standardized experimental settings.

Moreover, the diagnostic label of the participants in our sample was partly influenced by the results of ADOS and ADI-R. Thus, while the follow-up correlation analyses might shed light on underlying commonalities in autistic symptomatology between participants in our classification, they are not conclusive.

Finally, and importantly, even though we have implemented a careful and rather conservative cross-validation structure within our model, the sample size in this study is limited, and the results require external validation. As this study served as a proof-of-concept, the present videos were chosen based on a meticulous screening process, which consequently resulted in a high number of exclusions. For example, we only analyzed video excerpts of more than five minute in length and without the use of any external props; the latter of which is an important part of the ADOS assessment. However, we are confident that the high scalability of the methodology used in this study will encourage future data collection and, hence, further external and cross-site validation. In this regard, it will be important to analyze any effects of relaxed inclusion criteria concerning, e.g., the minimum length of an analysis window for a feasible synchrony assessment.

While clinicians’ judgments continue to outperform computational algorithms in their diagnostic precision [51], the notion of digital augmentation of the diagnostic process could prospectively loosen the current bottlenecks caused by resource-exhaustive clinical assessments. Considering the aforementioned limitations, we present a viable route toward a digitally assisted diagnostic process in autism. Using a heterogeneous dataset, both in age and technical setup, our classification model could detect ASD in a clinical sample with an above-chance accuracy. With few adjustments regarding the standardization of the experimental setup, including possibilities to record nuanced facial expression and vocal output, the strength of our approach has the potential for high scalability in everyday clinical practice.

## 6 Acknowledgements

We thank Afton Nelson for proofreading this manuscript.

## 7 Authors’ contribution

HY and CFW conceptualized the study. JCK and CFW designed the pre-processing procedure. DS and GB compiled and pre-processed the data. JCK and MSD analyzed and interpreted the data. NK supervised the machine learning analysis. JCK wrote the manuscript. All authors read and approved the final manuscript.

## 8 Availability of data and materials

The datasets generated or analyzed during the study are not publicly available as the IRB approved the data to be used within the research team but could be available from the corresponding author on reasonable request. The preprocessing scripts used during this study are available under https://github.com/jckoe/SNU_ASDsync.git

