## Supplementary Information for "Classifying autism in a clinical population based on motion synchrony: a proof-of-concept study using real-life diagnostic interviews"

^1^Department of Psychiatry and Psychotherapy, LMU University Hospital, LMU Munich, Germany, ^2^Department of Psychiatry, Seoul National University Bundang Hospital, Seongnam, South Korea, ^3^Department of Psychiatry, Seoul National University College of Medicine, Seoul, Korea, ^4^Max Planck Institute of Psychiatry, Munich, Germany, ^5^Institute of Psychiatry, Psychology and Neuroscience, King’s College, London, United Kingdom

*^*^equally contributing first authors*

*^+^equally contributing senior authors*

Corresponding author:

Jana C. Koehler,

LMU Clinic for Psychiatry and Psychotherapy

Nussbaumstr. 7

80336 Munich

+49 89 4400 55336

### S2 Supplementary methods

#### S2.1 Vocal Output Analysis

Audio tracks of the selected video clips were annotated within a semi-automated pipeline in the software Praat [1]. Given certain silence and sounding thresholds, the software automatically extracts a time series of silence and sounding intervals in an audio file. Minimum sounding and silent interval duration were set to 200 ms [2]. The minimum silence threshold was adjusted for each clip individually between 20-30 dB. The resulting intensity time series were checked visually for any noise artefacts caused by, e.g., paper shuffling or moving chairs. This way, it was ensured that the sounding intervals contained actual vocal output. Finally, the cleaned time series for each dyad were submitted to further analysis in RStudio [3].

Since the audio was captured via the camera the dyads were recorded with, individual channel separation was not employed. Consequently, in order to get an approximation of the verbal interaction for each dyad, we extracted a vocalization-to-silence ratio, which was calculated as the total vocal output duration (s) divided by the total silence duration. The distribution of this ratio across groups can be seen in Supplementary Figure 1.

Supplementary Figure 1. Vocalization-to-silence ratio per group

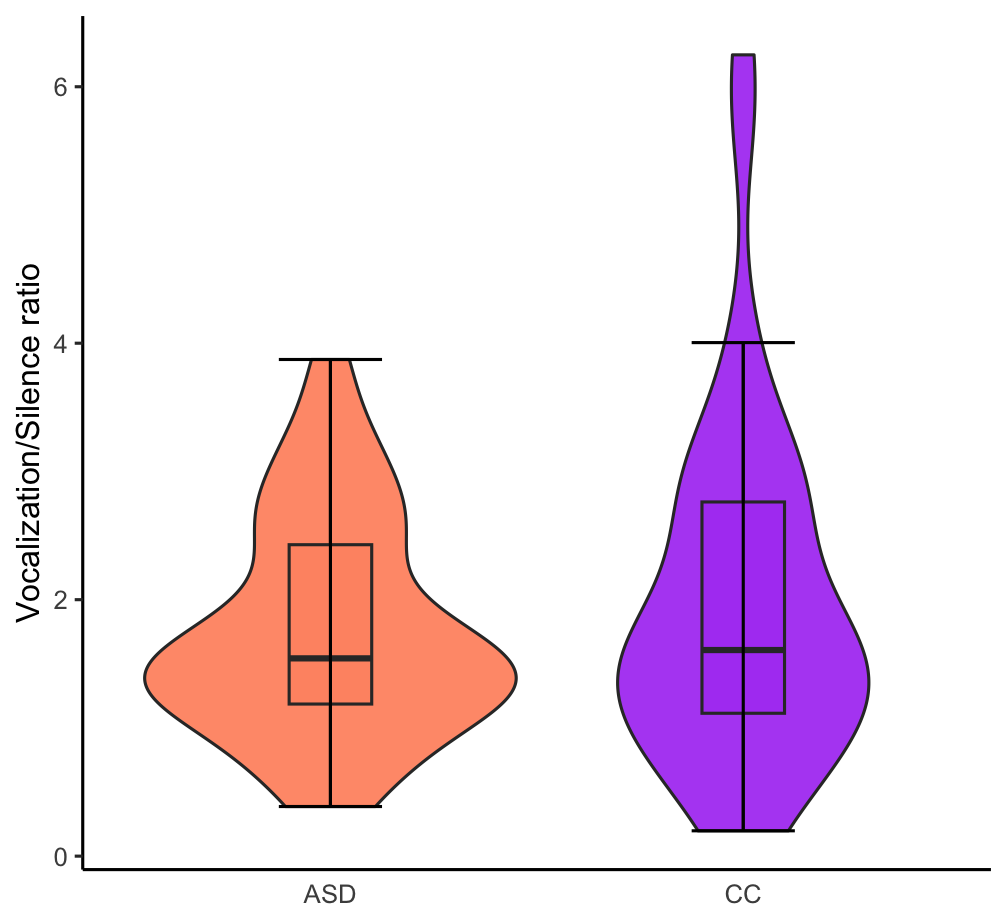

Note. Bars represent standard error.

#### S2.2 SVM: Predictive pattern extraction methods

In addition to the conventional feature weight extraction, we computed two other measures to visualize the predictive patterns of the features in our model: the cross-validation ratio (CVR, Supplementary Figure 2) and sign-based consistency (Supplementary Figure 3).

##### S2.2.1 Cross-validation ratio (CVR)

As a measure of stability, we calculated the cross-validation ratio (CVR), which was defined as the sum across CV2 folds of all CV1 median weights divided by the CV1 standard error, subsequently divided by the number of CV2 folds. This was based on the bootstrap ratio, a measure for pattern stability commonly used in the Partial Least Squares literature [4]. Similarly to the bootstrap ratio, the CVR of pattern element 𝑗, be it a statistical estimate of the inter- or intrapersonal synchrony, or a demographic variable, was defined as:

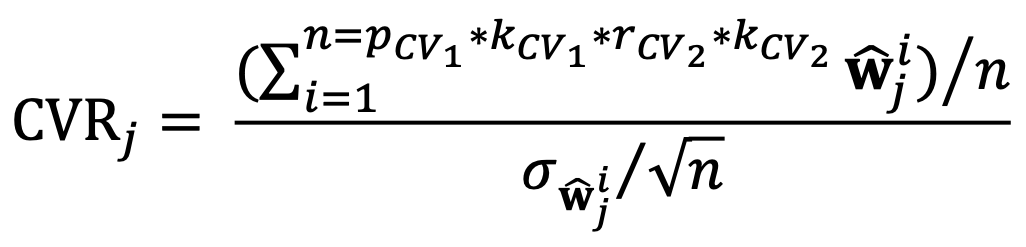
where 𝑛 is the size of the SVM ensemble, 𝑝_𝐶𝑉1_ is the number of CV1 permutations, 𝑘_𝐶𝑉1_ the number of CV1 folds, 𝑟_𝐶𝑉2_ the number of CV2 repetitions, 𝑘_𝐶𝑉2_ the number of CV2 folds, 𝐰̂^𝑖^_j_ the 𝑗th element of 𝑖th normalized weight vector 𝐰̂^𝑖^ = 𝐰^𝑖^/‖𝐰^𝑖^‖ in the SVM ensemble, 𝜎𝐰̂^𝑖^_j_ the standard deviation of 𝐰̂^𝑖^_j_. Similar to *Z*-scores, the CVR vectors were thresholded at CVR value ranges corresponding to an alpha level of 0.01 to delineate stable pattern elements across the cross-validation experiment.

Supplementary Figure 2. Cross-validation ratio of all features for SVM model based on motion energy synchrony.

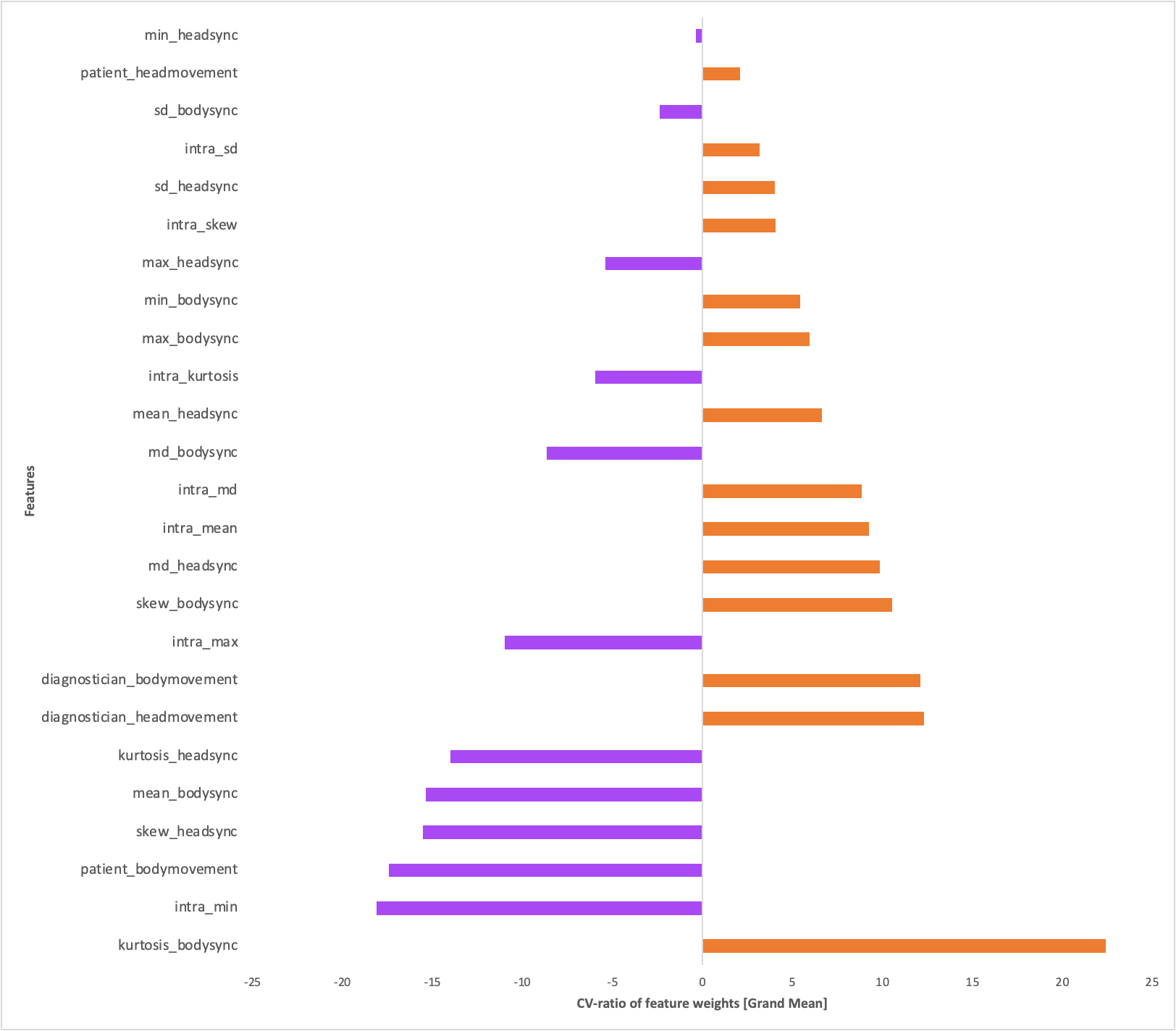

Note. Figure depicts the sum across CV2 folds of the selected CV1 median weights divided by the selected CV1 standard error, which is subsequently divided by the number of CV2 folds. Absolute values >= 2 correspond to p <= .05, absolute values >=3 correspond to p <= .01.

##### S2.2.2 Sign-based consistency

As a further measure of pattern stability, we used a sign-based consistency approach to statistically investigate the relevance of the variables in our SVM model. We based this method on the approach by Gómez-Verdejo and colleagues [5] toward wrapper-based feature selection strategies. The variable importance of the 𝑗th pattern element in the SVM model was defined as:

The first part of
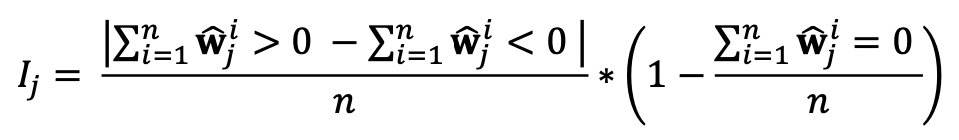
the equation measures the consistency of the weights assigned by the SVM ensemble to a given pattern element. The second part measures the fraction of SVMs that de-selected the given pattern element during the wrapper-based optimization process, which reduces the importance 𝐼*_j_*. Thus, 𝐼*_j_* = 1 represents perfect consistency as the weights of 𝑗th pattern element all share the same sign and the element has been selected by all classifiers in the ensemble. On the other hand, 𝐼*_j_* = 0 occurs when the weights are equally positive and negative across the ensemble or when the pattern element has been assigned zero weight by all classifiers in the ensemble. We defined a hypothesis test for 𝐼*_j_* , with:

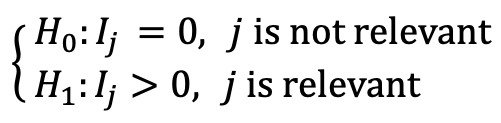
Subsequently, a z-score was calculated as follows:
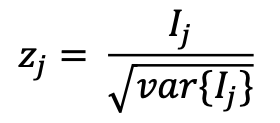

We then calculated a standard right-tailed p-value using a normal cumulative distribution function. Statistical significance was defined at α=0.05. The *p* values were then corrected for multiple comparisons using the false discovery rate.

Supplementary Figure 3. Sign-based consistency values of all features for SVM model based on motion energy synchrony.

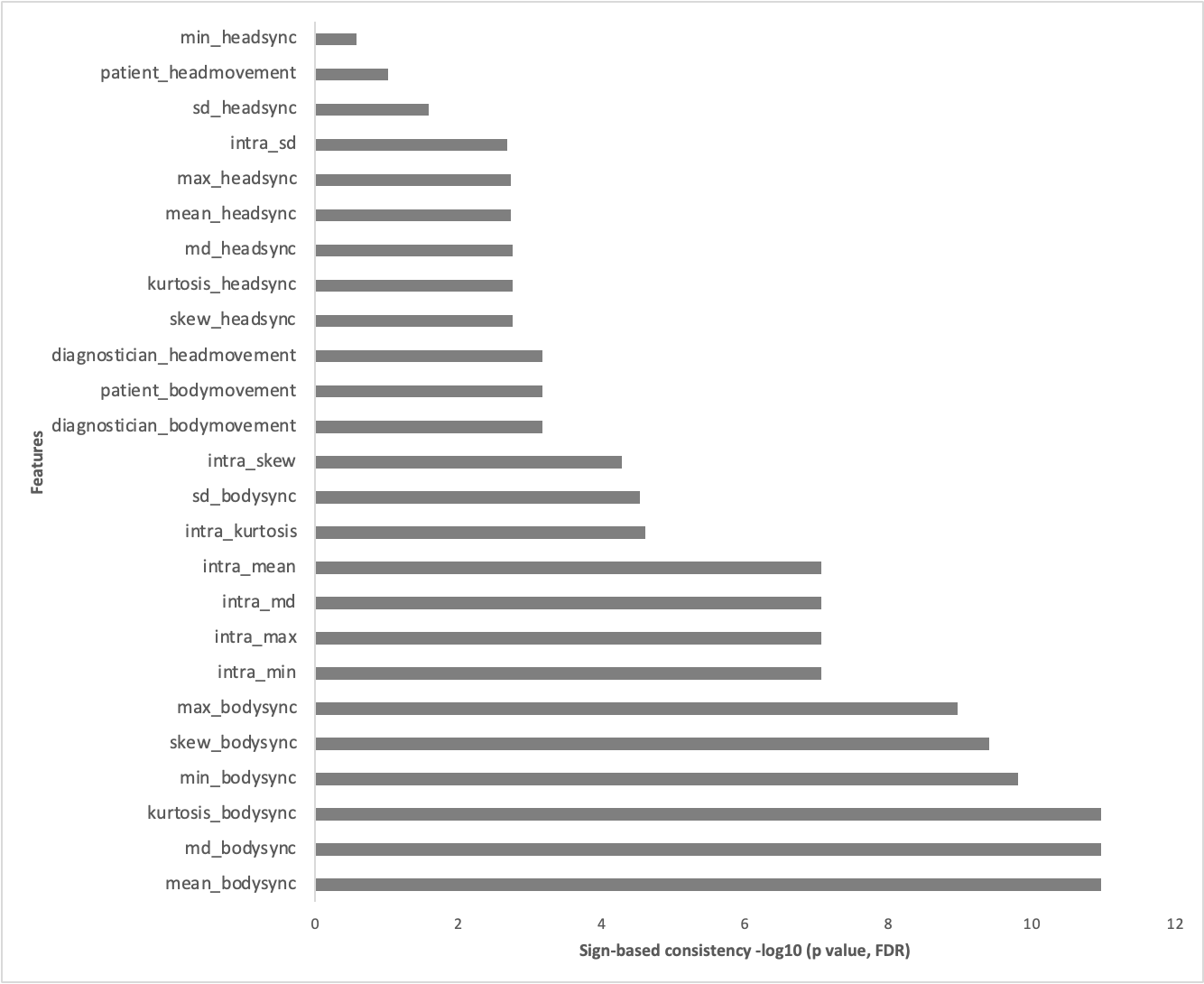

Note. The importance of each feature was calculated as the number of times that the sign of the feature was consistent. The depicted scores represent the resulting negative logarithm of p values that were corrected using the Bonferroni-Holm false-discovery rate. Sign-based consistency -10log(p) >= 1.3 is equivalent to p <= .05.

### S3 Supplementary results

#### S3.1 Differences in videos across groups

The videos used for this study differed in both length and conversational content. To rule out any classification bias caused by this, we compared video length and number of ADOS tasks depicted in each video across groups. No significant differences were found for either of the two variables.

| **Variables** | ***ASD*** | | **CC** | | **T test** | | | **Effect size** |
| --- | --- | --- | --- | --- | --- | --- | --- | --- |
|  | **M** | **SD** | **M** | **SD** | ***t*** | ***df*** | ***p_adj_*** |  |
| Video Length (s) | 437.95 | 112.76 | 443.82 | 11.84 | -.24 | 78.10 | .808 | -.05 |
| # of ADOS tasks included in video | 2.07 | .57 | 2.26 | .72 | -1.37 | 66.50 | .175 | -.30 |
| Note. p values corrected for multiple comparisons using Benjamini-Hochberg method. | | | | | | | | |

#### S3.2 Room differences

We investigated whether the differing video setup impacted our features. Using analyses of variance (ANOVAs), we compared the mean head, body and intrasync values between the different rooms. Results can be found in the table below.

Supplementary Table 1. Feature differences across rooms.

| Measure | Room 1  (n = 44) | | Room 2  (n = 38) | | Room 3  (n = 10) | | Room 4  (n = 2) | | F(3,90) | *p* |
| --- | --- | --- | --- | --- | --- | --- | --- | --- | --- | --- |
|  | M | SD | M | SD | M | SD | M | SD |  |  |
| Head Synchrony | .10 | .02 | .10 | .01 | .10 | .01 | .09 | .02 | .547 | .915 |
| Body Synchrony | .10 | .03 | .10 | .02 | .11 | .02 | .10 | .02 | .173 | .651 |
| Intrasynchrony | .21 | .05 | .21 | .04 | .22 | .04 | .02 | .00 | .155 | .927 |

Supplementary Figure 4. Feature means across different testing rooms.

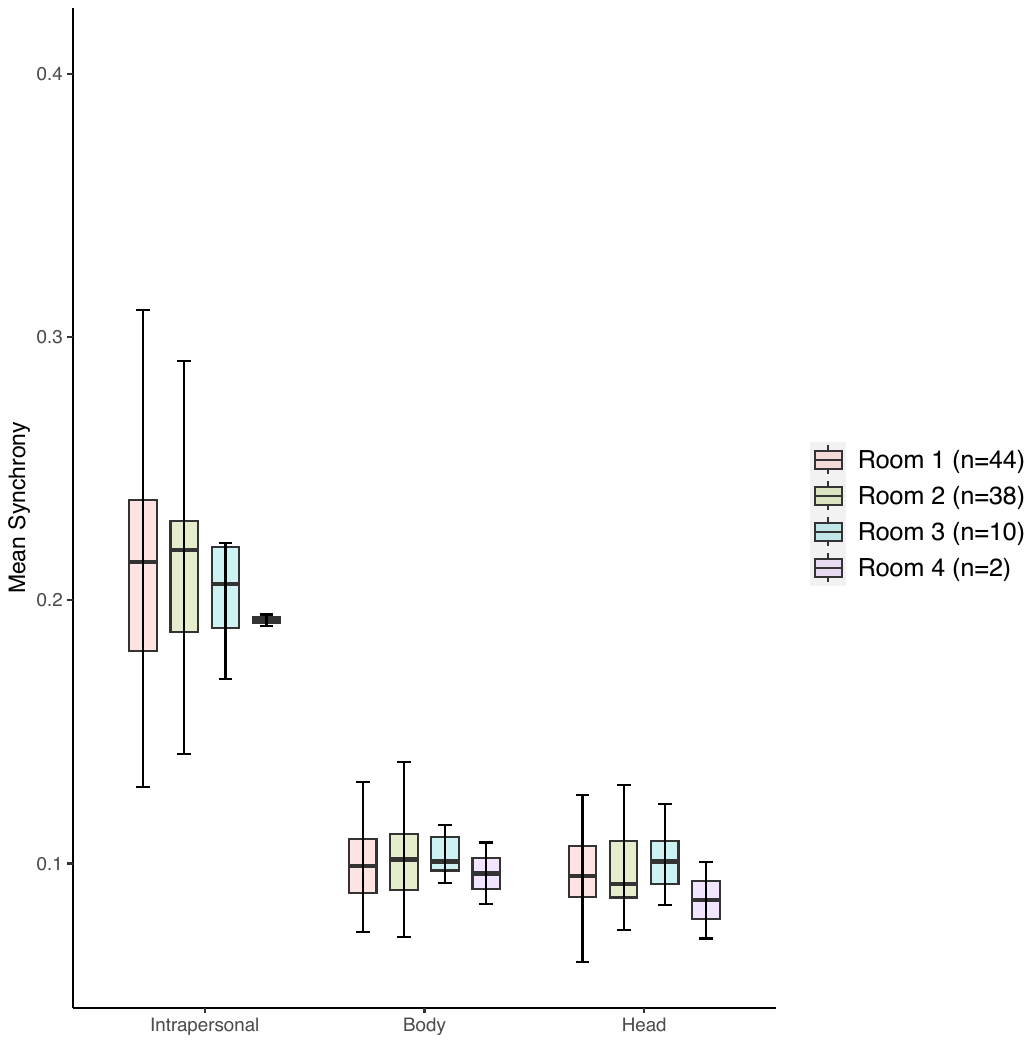

#### S3.3 Pseudosynchrony

Following procedures from previous MEA publications, a measure of pseudosynchrony was computed and compared to the interpersonal synchrony values extracted in our sample. Though both head and body synchrony were lower in the artificial pseudodyads both descriptively (Supplementary Table 2) and visually (Supplementary Figure 3), this difference was not significant.

Supplementary Table 2. Results of mean comparison of interpersonal synchrony in real and pseudo-dyads

|  | Synchrony | | Pseudosynchrony | | *t*(592) | *p* | Cohen’s *d* |
| --- | --- | --- | --- | --- | --- | --- | --- |
|  | *M* | *SD* | *M* | *SD* |  |  |  |
| Head | .098 | .016 | .096 | .017 | .841 | .401 | .095 |
| Body | .104 | .022 | .102 | .019 | .798 | .425 | .089 |
| Note. Results computed using two sample independent t-tests assuming equal variances. | | | | | | | |

Supplementary Figure 5. Density plots of average cross-correlations per group and ROI.

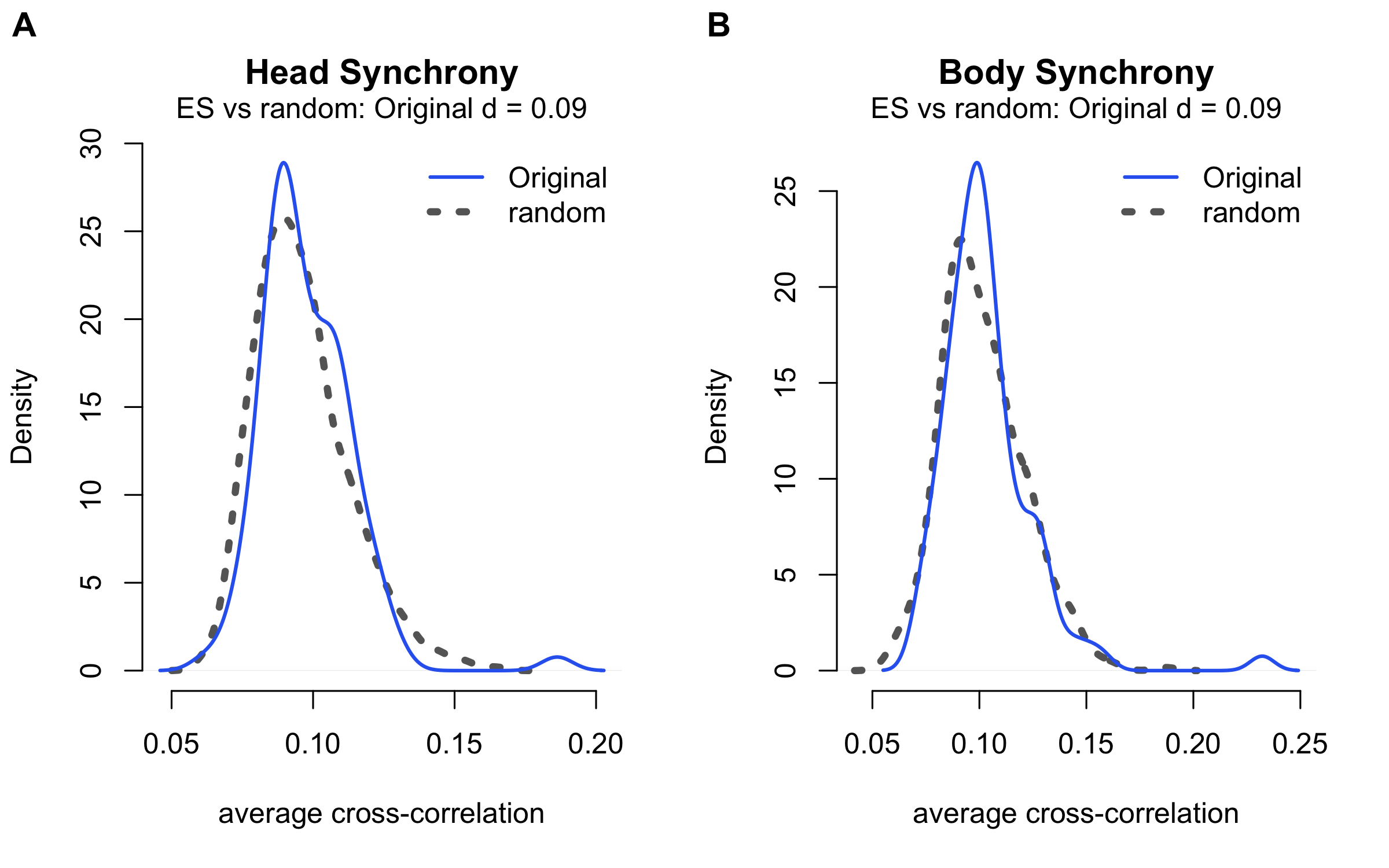
Note. Effect size computed using Cohen’s d.

#### S3.4 SVM model with features derived from peak-picking

In a previous study investigating naturalistic conversations between adult autistic and non-autistic participants [6], we operationalized interpersonal synchrony as the instances of maximum synchrony within the interaction, derived by a peak-picking algorithm [7], [8]. The same windowed cross-lagged correlation was run on the smoothed time series for interpersonal (window size of 60s, lags of 5s, increments of 30s) and intrapersonal synchrony (window size of 30s, lags of 5s, increments of 15s). The peak-picking procedure then picks the highest cross-correlation per window in every dyad. Subsequently, every dyad summary statistic (mean, median, minimum, maximum, standard deviation, skewness, kurtosis) of the peaks were computed and used as feature set in a corresponding SVM analysis, using the same parameter setup as the main analysis and regressing out age. The results are shown below (Supplementary Table 3).

Supplementary Table 3. SVM classification metrics of dyad classification using trajectory of peak synchrony instances.

| Model | BAC (%) | Sensitivity (%) | Specificity (%) | AUC | TN | TP | FN | FP | Accuracy (%) | Number needed to diagnose | Positive likelihood ratio | Diagnostic odds ratio | Permutation test, *p* value |
| --- | --- | --- | --- | --- | --- | --- | --- | --- | --- | --- | --- | --- | --- |
| ASD vs. CC | 53.2 | 64.3 | 42.1 | .55 | 16 | 36 | 20 | 22 | 55.3 | 15.6 | 1.1 | 1.2 | .166 |
| Note. ASD = Autism Spectrum Disorder. CC = Clinical Control. NDD = Neurodevelopmental Disorder. BAC = Balanced Accuracy. AUC = Area Under The Receiver Operating Curve. TN = True Negatives. TP = True Positives. FN = False Negatives. FP = False Positives. | | | | | | | | | | | | | |

### S4 Video setup instructions

For improved scalability and usability in computer vision analyses, we propose recommendations for a standardized video setup in ADOS interviews (Supplementary Figure 4).

Supplementary Figure 6. Instructions for the standardized taping of prospective ADOS interviews.

**[ADOS]**

1. To account for movement by the participant, try to capture a wide range (no zooming)
   *Keep the same camera angle and lighting (crucial for region of interest analysis)*

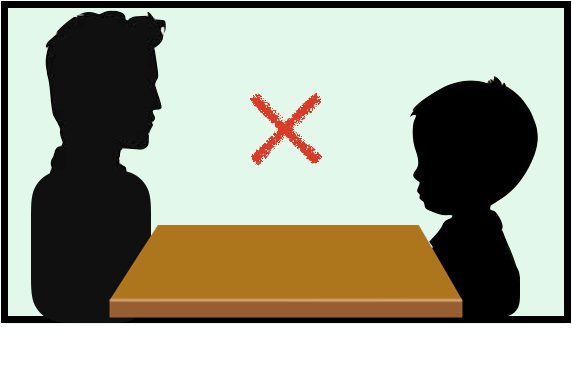

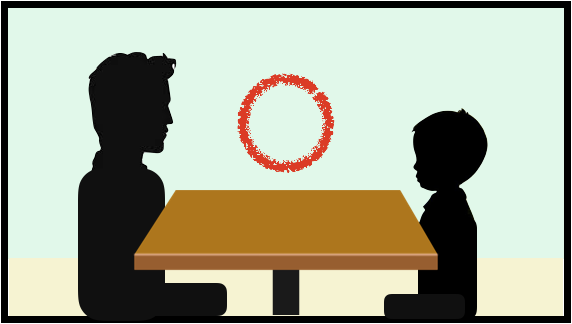

2. Make sure the camera is placed to capture the side angle (not diagonal)

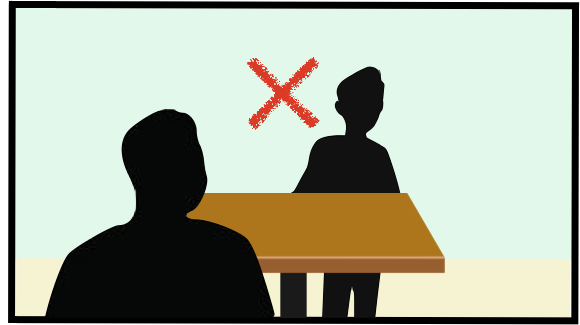

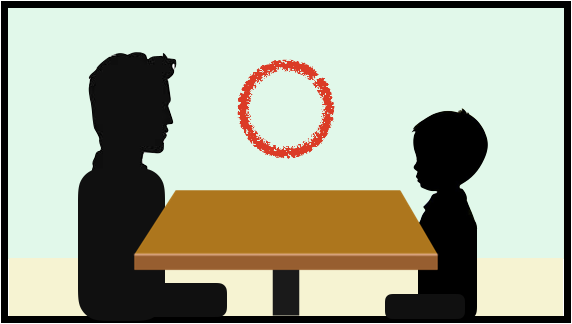

3. Check to see if the upper body of the administrator or participant is cut off

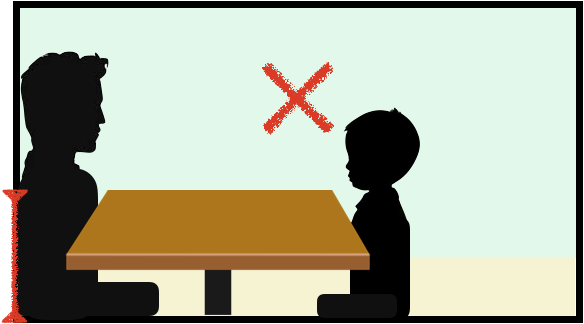

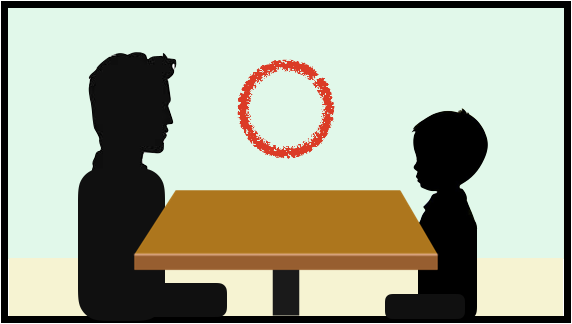

4. Check to see if the head of the administrator or participant is cut off

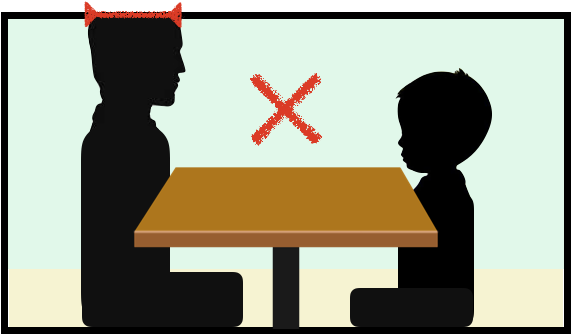

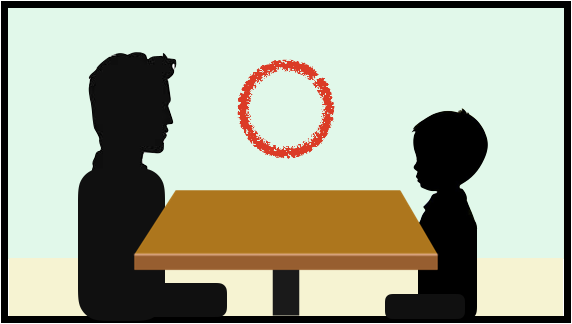

5. Check to see if there are any objects that may be covering the administrator or participant

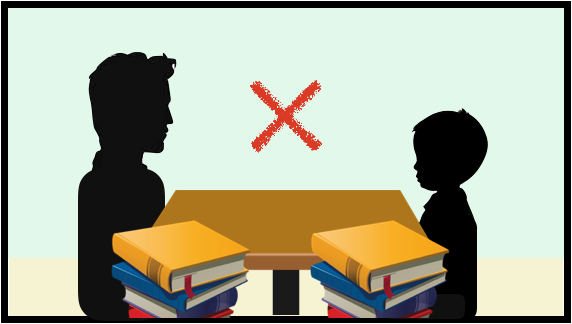

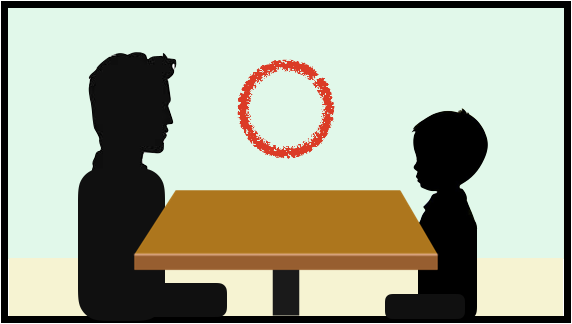

6. Make sure there are no fingerprints on the camera lens

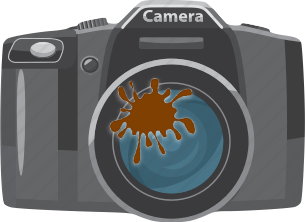

** See if it is possible to replace the chair for the participant with something that is more fixed or stable. As the current chair is not fixed, a number of children who were swinging in their chairs.

Note. A Korean translation of these recommendations are available from the corresponding author upon request.

### S5 Additional supplementary tables

#### S5.1 Supplementary Table. Additional medical information on study subsample

| **Psychopathological profile** |  | ASD (n = 19) | CC (n = 20) |
| --- | --- | --- | --- |
|  | ADHD | 4 | 10 |
|  | Bipolar Disorder | - | 2 |
|  | Depressive Disorder | 1 | 4 |
|  | Intellectual Disorder | - | 4 |
|  | Anxiety Disorder | 1 | 1 |
|  | Broad Spectrum | - | 2 |
|  | Developmental Delay | - | 1 |
|  | Social Anxiety | - | 1 |
|  | Tourette Syndrome | - | 1 |
|  | Tic Disorder | 1 | - |
|  | OCD | 1 | - |
|  | Communication Disorder | 1 | - |
| **Medication** |  |  |  |
|  | Anticonvulsant | 1 | 3 |
|  | Antidementive | 1 | - |
|  | Antidepressant | 7 | 10 |
|  | Antipsychotic | 6 | 6 |
|  | Antispasmodic | - | 1 |
|  | Benzodiazepine | 2 | 3 |
|  | Beta-Blocker | 1 | 1 |
|  | Nonbenzodiazepine | - | 1 |
|  | Psychostimulant | 3 | 1 |
|  | Others | - | 2 |
| Note. Information on comorbidities and medication is part of the standardized procedure within the clinical assessment in the outpatient clinic at the study site and is therefore only available for this subsample. | | | |

#### S5.2 Supplementary Table. Full feature set for SVM model

| Modality | Features |
| --- | --- |
| Interpersonal Head Synchrony – Full cross-correlation matrix | min_headsync  max_headsync  mean_headsync  md_headsync  sd_headsync  skew_headsync  kurtosis_headsync |
| Interpersonal Body Synchrony – Full cross-correlation matrix | min_bodysync  max_bodysync  mean_bodysync  md_bodysync  sd_bodysync  skew_bodysync  kurtosis_bodysync |
| Intrapersonal Head-Body Coordination within patient– Full cross-correlation matrix | intra_min  intra_max  intra_mean  intra_md  intra_sd  intra_skew  intra_kurtosis |
| Total amount of movement for patient and administrator | patient_headmovement  patient_bodymovement  diagnostician_headmovement  diagnostician_bodymovement |
| *Note.* min = mimimum, max = maximum, md = median, sd = standard deviation, skew = skewness | |

#### S5.3 Supplementary Table. Detailed classification metrics for SVM model based on Motion Energy Synchrony Analyses and demographic features

| Model | BAC (%) | Sensitivity (%) | Specificity (%) | AUC | TN | TP | FN | FP | Accuracy (%) | Number needed to diagnose | Positive likelihood ratio | Diagnostic odds ratio | Permutation test, *p* value |
| --- | --- | --- | --- | --- | --- | --- | --- | --- | --- | --- | --- | --- | --- |
| ASD vs. CC | 59.4 | 71.4 | 47.4 | .58 | 18 | 40 | 16 | 20 | 61.7 | 5.3 | 1.4 | 1.8 | .017 |
| Note. ASD = Autism Spectrum Disorder. CC = Clinical Control. NDD = Neurodevelopmental Disorder. BAC = Balanced Accuracy. AUC = Area Under The Receiver Operating Curve. TN = True Negatives. TP = True Positives. FN = False Negatives. FP = False Positives. | | | | | | | | | | | | | |

#### S5.4 Supplementary Table. Pearson correlation coefficients for the association between the MEA model SVM decision scores and clinical variables.

|  | n | M | SD | 1 | *P_adj_* |
| --- | --- | --- | --- | --- | --- |
| 1. SVM decision score | 94 | .19 | .89 | - | - |
| 2. ADI-R_A | 84 | 12.67 | 8.10 | .07  [-.15, .28] | .662 |
| 3. ADI-R_B | 84 | 9.33 | 5.79 | .07  [-.15, .28] | .662 |
| 4. ADI-R_C | 84 | 3.46 | 2.80 | .16  [-.06, .36] | .662 |
| 5. ADOS_RRB_CSS | 94 | 3.73 | 2.93 | .08  [-.12, .28] | .662 |
| 6. ADOS_SA_CSS | 94 | 6.03 | 2.53 | .02  [-.19, .22] | .894 |
| 7. ADOS_Total_CSS | 94 | 5.32 | 2.58 | .01  [-.19, .22] | .894 |
| Note. ADOS scores were transformed to calibrated severity scores as described in [9], [10]. p values adjusted for multiple comparisons using Benjamini-Hochberg FDR-method. M and SD are used to represent mean and standard deviation, respectively. Values in square brackets indicate the 95% confidence interval. | | | | | |
